# Insights into Ancestral Diversity in Parkinson’s Disease Risk: A Comparative Assessment of Polygenic Risk Scores

**DOI:** 10.1101/2023.11.28.23299090

**Authors:** Paula Saffie-Awad, Mary B Makarious, Inas Elsayed, Arinola O. Sanyaolu, Peter Wild Crea, Artur F Schumacher Schuh, Kristin S Levine, Dan Vitale, Mathew J Koretsky, Jeffrey Kim, Thiago Peixoto Leal, María Teresa Periñan, Sumit Dey, Alastair J Noyce, Armando Reyes-Palomares, Noela Rodriguez-Losada, Jia Nee Foo, Wael Mohamed, Karl Heilbron, Lucy Norcliffe-Kaufmann, the 23andMe Research Team, Mie Rizig, Njideka Okubadejo, Mike A Nalls, Cornelis Blauwendraat, Andrew Singleton, Hampton Leonard, Ignacio F. Mata, Sara Bandres-Ciga, the Global Parkinson’s Genetics Program (GP2)

## Abstract

**Objectives:** To evaluate and compare different polygenic risk score (PRS) models in predicting Parkinson’s disease (PD) across diverse ancestries, focusing on identifying the most suitable approach for each population and potentially contributing to equitable advancements in precision medicine.

**Methods:** We constructed a total of 105 PRS across individual level data from seven diverse ancestries. First, a cross-ancestry conventional PRS comparison was implemented by utilizing the 90 known European risk loci with weighted effects from four independent summary statistics including European, East Asian, Latino/Admixed American, and African/Admixed. These models were adjusted by sex, age, and principal components (28 PRS) and by sex, age, and percentage of admixture (28 PRS) for comparison. Secondly, a novel and refined multi-ancestry best-fit PRS approach was then applied across the seven ancestries by leveraging multi-ancestry meta-analyzed summary statistics and using a p-value thresholding approach (49 PRS) to enhance prediction applicability in a global setting.

**Results:** European-based PRS models predicted disease status across all ancestries to differing degrees of accuracy. Ashkenazi Jewish had the highest Odds Ratio (OR): 1.96 (95% CI: 1.69-2.25, p < 0.0001) with an AUC (Area Under the Curve) of 68%. Conversely, the East Asian population, despite having fewer predictive variants (84 out of 90), had an OR of 1.37 (95% CI: 1.32-1.42) and an AUC of 62%, illustrating the cross-ancestry transferability of this model. Lower OR alongside broader confidence intervals were observed in other populations, including Africans (OR =1.38, 95% CI: 1.12-1.63, p=0.001). Adjustment by percentage of admixture did not outperform principal components. Multi-ancestry best-fit PRS models improved risk prediction in European, Ashkenazi Jewish, and African ancestries, yet didn’t surpass conventional PRS in admixed populations such as Latino/American admixed and African admixed populations.

**Interpretation:** The present study represents a novel and comprehensive assessment of PRS performance across seven ancestries in PD, highlighting the inadequacy of a ‘one size fits all’ approach in genetic risk prediction. We demonstrated that European based PD PRS models are partially transferable to other ancestries and could be improved by a novel best-fit multi-ancestry PRS, especially in non-admixed populations.

## INTRODUCTION

The heritability attributed to idiopathic Parkinson’s disease (PD) in European populations is estimated to be around 22%^1^. Genome-wide association studies (GWAS) have been key at identifying common loci that contribute to PD risk. A total of 90 risk variants across 78 independent loci have been associated with PD risk in European ancestry populations^1^. More recently, large-scale efforts are focusing on increasing genetic diversity in PD studies to unravel the genetic architecture of disease across ancestries^2–5^. The first and largest trans-ethnic PD GWAS meta-analysis performed to date in European, East Asian, Latino/Admixed American, and African ancestry populations identified a total of 78 loci reaching or maintaining genome-wide significance, 12 of which had not been previously identified^6^.

A polygenic risk score (PRS) can be generated to estimate an individual’s susceptibility to a binary outcome, exploring the cumulative estimated effect of common genetic variants on an individual’s phenotype like PD^7,8^. In this context, PRS alone has not been shown to have clinical utility in predicting PD in European populations, with only 56.9% sensitivity and 63.2% specificity to predict disease at best^9^. PRS utility improves both sensitivity (83.4%) and specificity (90.3%) when including relevant clinical criteria such as olfactory function, family history, age, and gender^9,10^. Similarly, the integration of environmental factors ameliorates case/control stratification ^10,11^ while the combination of multi-omics and clinical criteria in PRS models boosts prediction across multiple diseases ^11,12^.

Nevertheless, the current focus on European ancestries in PRS development highlights a significant research gap. Using PRS to calculate disease risk in a single population may exacerbate existing health disparities as it cannot be accurately implemented across diverse ancestries ^13,14^. Despite challenges in the direct applicability of European-ancestry PRS, there’s growing evidence for their cross-ancestry transferability, as seen in their association with diseases like Alzheimer’s^15^, breast cancer^16^, and venous thromboembolism among non-European groups^17^. This indicates potential for methodological refinements to improve PRS reliability and address health equity concerns^18–20^.

In the PD genetics field, studies investigating how cumulative genetic risk varies within and between different ancestral populations have not been conducted. Here, we perform the most comprehensive assessment of PRS in PD by implementing two approaches: First, we explore differences in the application and generalizability of the conventional PD PRS model using population-specific summary statistics across seven individual-level cohorts of diverse ancestry populations, including East Asians, Central Asians, Latino/Admixed American, Africans, African admixed, and Ashkenazi Jewish individuals. Secondly, we build multi-ancestry best-fit PRS models for these diverse ancestry populations based on summary statistics from a recent PD multi-ancestry GWAS meta-analysis^21^. By doing so, we aim to provide insights that will lead to the development of more accurate and inclusive genetic prediction models for PD research, thereby enhancing PRS’s predictive power and contributing to equitable advancements in precision medicine.

## METHODS

### Study Participants

Our study workflow is highlighted in **Figure 1**. We obtained multi-ancestry individual-level data from the Global Parkinson’s Genetics Program (GP2) https://gp2.org/^22^ release 6 (doi: 10.5281/zenodo.10472143, https://doi.org/10.5281/zenodo.10472143. These data (here referred to as ***target data***) were used to test PRS models and comprised a total of 41,831 participants, including 24,709 individuals diagnosed with PD, 17,246 controls, and 2,876 participants diagnosed with neurological diseases other than PD. After excluding related individuals (those at the first cousin level or closer) that could bias our PRS assessments and those classified as non-PD cases, our dataset comprised a total of 29,051 individuals, of which 15,989 were PD cases and 13,062 controls. The following genetic ancestries were included: African admixed, African, Ashkenazi Jewish, Latino/Admixed American, Central Asian, East Asian, and European populations (**Supplementary Figure 1**, see Methods for ancestry clustering description). Detailed demographic and clinical characteristics can be found in **Table 1**.

**Figure 1:**
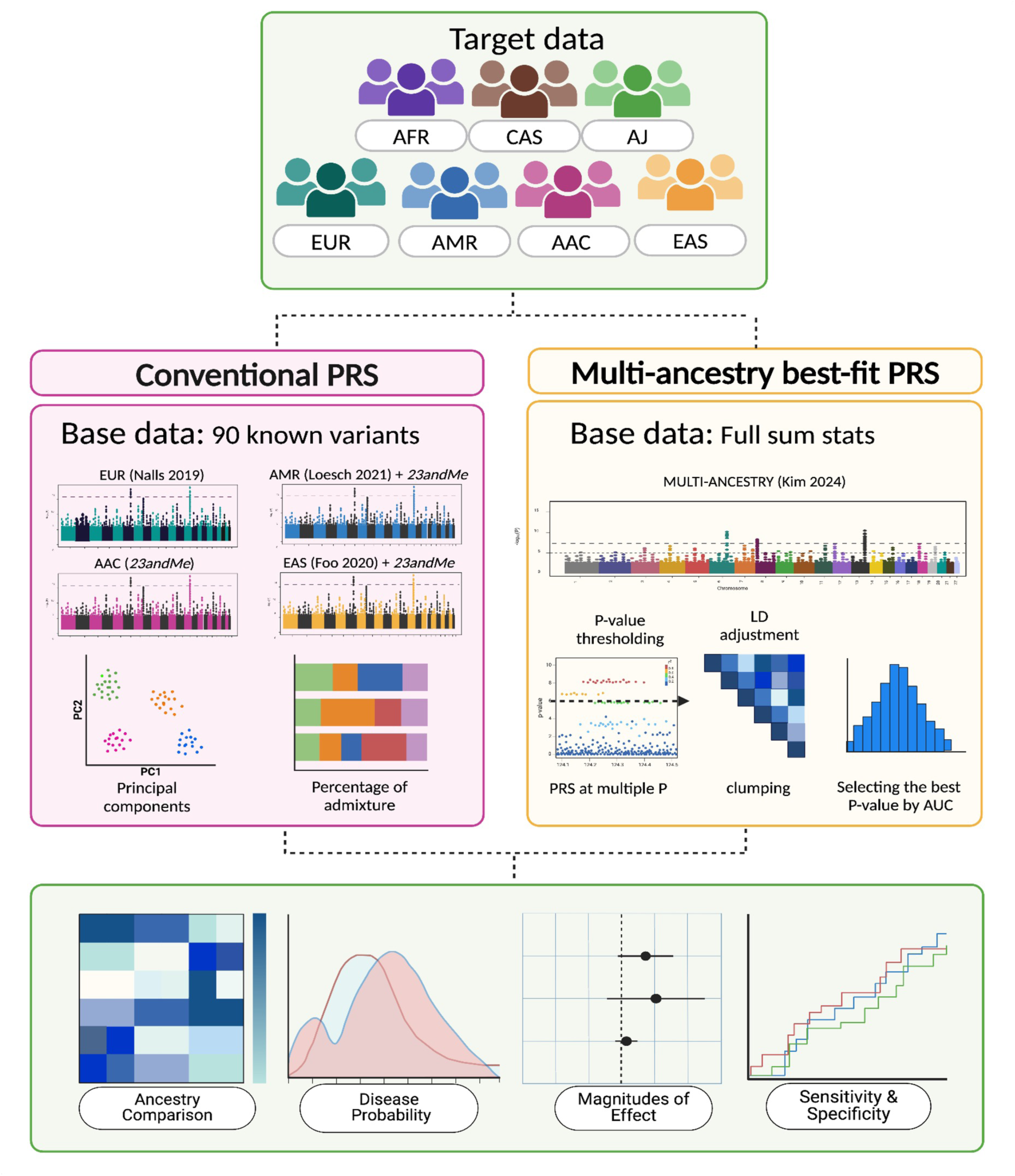
Schematic study workflow Summarized study diagram divided in three panels. The first panel displays the target data, from seven diverse ancestry groups: African Admixed (AAC), African (AFR), Ashkenazi Jewish (AJ), Latino/Admixed American (AMR), Central Asian (CAS), East Asian (EAS), and European (EUR). In the second panel, the two models being compared are detailed: a) The Conventional PRS approach, which evaluates 90 PD risk variants identified by Nalls et al., 2019, in each of the seven ancestries weighted by the effect sizes derived from four population-specific GWAS (EUR, AAC, AMR, EAS) and adjusted by principal components and percentage of admixture, leading to the generation of 56 scores; b) The Multi Ancestry Best Fit PRS approach, which employs the PRSice software tool, computing p-value thresholding along variant-specific weights by leveraging full summary statistics from Kim et al., 2024 (pruned using default parameters). The outcomes are visualized in the third panel, through heatmaps for ancestry comparison, density plots for disease probability, forest plots for effect size, and Receiver Operating Characteristic (ROC) plots for evaluating the models’ sensitivity and specificity.

**Table 1.** Demographic characteristics of individual level data and power calculation.

| Ancestry | Total | Male (n, %) | Cases |  | Controls |  | Power |
| --- | --- | --- | --- | --- | --- | --- | --- |
|  |  |  | n | AAO (mean ± SD) | n | Age (mean ± SD) |  |
| EUR | 19138 | 12541 (61.23%) | 11656 | 57.95 ± 12.00 | 7482 | 65.03 ± 10.56 | 100.00% |
| AAC | 1058 | 439 (40.92%) | 255 | 58.57 ± 12.58 | 803 | 65.84 ± 10.43 | 81.99% |
| AMR | 502 | 286 (54.98%) | 363 | 50.19 ± 13.88 | 139 | 63.17 ± 9.04 | 30.09% |
| EAS | 3827 | 2510 (65.01%) | 1534 | 49.85 ± 12.87 | 2293 | 62.37 ± 11.21 | 99.98% |
| AFR | 2539 | 1430 (56.12%) | 916 | 57.24 ± 12.85 | 1623 | 63.55 ± 15.40 | 99.51% |
| AJ | 1458 | 1321 (68.51%) | 1001 | 60.26 ± 11.61 | 457 | 67.77 ± 9.66 | 73.13% |
| CAS | 529 | 264 (49.15%) | 264 | 47.01 ± 13.56 | 265 | 55.14 ± 5.32 | 49.21% |

### Target data

#### Genotype data generation and quality control

We performed genotype data generation according to standard protocols from the Global Parkinson’s Genetics Program (GP2) https://gp2.org/ ^22^ release 6 (doi: 10.5281/zenodo.10472143, https://doi.org/10.5281/zenodo.10472143). In summary, samples were genotyped on the NeuroBooster array (v.1.0, Illumina, San Diego, CA) that includes 1,914,935 variants encompassing ancestry informative markers, markers for identity by descent determination, and X-chromosome SNPs for sex determination. Additionally, the array includes 96,517 customized variants. Automated genotype data processing was conducted on GenoTools ^23^, a Python pipeline built for quality control and ancestry estimation of data. Additional details can be found at https://pypi.org/project/the-real-genotools/ ^23^.

Quality control (QC) was conducted following standard protocols, with adjustments made to enhance precision and reliability. Samples exhibiting a genotype call rate below 98% (--mind 0.02), discordant sex determinations (0.25 <= sex F <= 0.75), or significant heterozygosity (F <= -0.25 or F >= 0.25) were excluded from the analysis. Additional QC measures involved the exclusion of SNPs with a missingness rate above 2%, variants deviating significantly from Hardy-Weinberg Equilibrium (HWE P value < 1E-4), and variants showing non-random missingness by case-control status (P≤1E-4) or by haplotype (P≤1E-4 per ancestry).

#### Ancestry predictions

Ancestry predictions were refined using an updated and expanded reference panel, which, as of January 2024, comprises samples from the 1000 Genomes Project (https://www.internationalgenome.org/data-portal/data-collection/phase-1)^24^, Human Genome Diversity Project^25^, and an Ashkenazi Jewish population dataset^26^. This panel includes 819 African, 74 African Admixed and Caribbean, 471 Ashkenazi Jewish, 183 Central Asian, 585 East Asian, 534 European, 99 Finnish, 490 Latino/Admixed American, 152 Middle Eastern, and 601 South Asian individuals. Palindromic SNPs were excluded to improve accuracy (AT or TA or GC or CG). Further filtering of SNPs within the panel was conducted to exclude variants with a minor allele frequency (MAF) below 0.05, a genotyping call rate less than 0.98, and Hardy-Weinberg equilibrium (HWE) p-value less than 1E-4. The process ensured the extraction of variants overlapping between the reference panel SNP set and the samples under study, totaling approximately 39,302 variants for ancestry estimations. Missing genotypes were imputed using the mean value of the variant from the reference panel.

To evaluate the efficacy of ancestry estimation, an 80/20 train/test split was applied to the reference panel samples, and principal components (PCs) were calculated using the overlapping SNPs. By applying transformations through UMAP, the global genetic population substructure and stochastic variation were visualized. Training a linear support vector classifier on the UMAP-transformed PCs resulted in consistent predictions, with balanced accuracies between 95% and 98%, as verified by 5-fold cross-validation on the test data from the reference panel. These classifier models were then applied to the dataset to generate ancestry estimates for all samples. Detailed methodologies for the cloud-based and scalable pipeline employed for genotype calling, QC, and ancestry estimation are documented in the GenoTools ^23^ GitHub repository (https://doi.org/10.5281/zenodo.10719034)^27^.

Following ancestry estimation, we excluded those with second-degree or closer relatedness (kinship coefficient > 0.0884). PCs that were used as covariates in the PRS analysis were recalculated per ancestry post-QC and ancestry determination. The percentage of ancestry was then computed using the supervised functionality of ADMIXTURE (v1.3.0; https://dalexander.github.io/admixture/binaries/admixture_linux-1.3.0.tar.gz), leveraging the labeled reference panel data to estimate ancestry proportions accurately.

#### Imputation

Variants with a minor allele frequency (MAF) of less than 0.05 and Hardy-Weinberg equilibrium (HWE) p-value less than 1E-5 were excluded before submission to the TOPMed Imputation server. The utilized TOPMed reference panel version, known as r2, encompasses genetic information from 97,256 reference samples and over 300 million genetic variants across the 22 autosomes and the X chromosome. As of October 2023, the TOPMed panel includes approximately 180,000 participants, with 29% of African, 19% of Latino/Admixed American ancestry, 8% of Asian ancestry, and 40% of European ancestry (https://topmed.nhlbi.nih.gov/). Further details about the TOPMed Study^28^, Imputation Server^29^, and Minimac Imputation^30^ can be accessed at https://imputation.biodatacatalyst.nhlbi.nih.gov. Following imputation, the resulting files underwent pruning based on a minor allele count (MAC) threshold of 10 and an imputation Rsq value of 0.3.

### Model 1: Conventional polygenic risk score approach

#### Ancestry-specific summary statistics generation

A total of four population-specific summary statistics were used to compute PRS versus the seven GP2 individual level data ancestry cohorts (**Supplementary Table 1a**). We obtained summary statistics for the European population from the largest European PD GWAS meta-analysis to date conducted by Nalls and colleagues (2019) (https://pdgenetics.org/resources). This study included 1,456,306 individuals, of which 1,400,000 were controls, 37,688 were cases and 18,618 were proxy cases (defined as having a first degree relative with PD). African admixed summary statistics were obtained from *23andMe*, which are based on 194,273 individuals including 193,985 controls and 288 cases. In order to achieve better-powered summary statistics for the East Asian population, we meta-analyzed two independent summary statistics including the largest East Asian PD GWAS meta-analysis to date^2^ and *23andMe* summary statistics from East Asian ancestry, which yielded a total of 183,802 individuals, including 176,756 controls and 7,046 cases. In a similar way, we conducted GWAS meta-analysis to generate better powered Latino/Admixed American summary statistics, combining the largest Latino PD GWAS meta-analysis from the LARGE-PD Consortium^3^ with *23andMe* Latino/Admixed American summary statistics. This cohort consisted of a total of 584,660 individuals, where 582,220 were controls and 2,440 PD cases. A summary of these data could be found in **Supplementary Table 1a.** *23andMe* participants provided informed consent and volunteered to participate in the research online, under a protocol approved by the external AAHRPP-accredited IRB, Ethical & Independent (E&I) Review Services. As of 2022, E&I Review Services is part of Salus IRB (https://www.versiticlinicaltrials.org/salusirb). The full GWAS summary statistics for the *23andMe* discovery data set will be made available through *23andMe* to qualified researchers under an agreement with *23andMe* that protects the privacy of the *23andMe* participants. Datasets will be made available at no cost for academic use. Please visit https://research.23andme.com/collaborate/#dataset-access/ for more information and to apply to access the data.

A comprehensive explanation of each step to generate *23andMe* summary statistics can be found elsewhere^31^. Briefly, the *23andMe* data generation process could be summarized in the following steps. After genotyping of *23andMe* participants was completed, an ancestry classifier algorithm was used to determine participant ancestries based on local ancestry and reference populations. Next, phasing was performed to reconstruct haplotypes using genotyping platform-specific panels followed by imputation of missing genotypes, expanding the variant dataset using two independent reference panels. Related individuals were then excluded using a segmental identity-by-descent estimation algorithm to ensure unrelated participants. Finally, a GWAS analysis adjusted by covariates age, sex, and principal components was conducted followed by GWAS QC measures to flag potential issues with SNPs, ensuring data integrity.

For a detailed description of the methods used to generate East Asian summary statistics, refer to the study by Foo et al.^2^. Similarly, detailed information on the Latino/Admixed American summary statistics can be found in Loesch et al.^3^. The GWAS meta-analysis of each population was carried out using fixed effects based on beta and SE values for the 90 risk variants. This meta-analysis was conducted utilizing the METAL package, which is accessible at https://genome.sph.umich.edu/wiki/METAL_Documentation.

#### Conventional polygenic risk score calculation

For conventional PRS calculations, we extracted the 90 risk predictors previously linked to PD risk in European ancestry populations^1^ from GP2 individual level data for each of the seven ancestries. Scores were weighted by the effect sizes derived from the four population-specific summary statistics previously mentioned (European, African Admixed, Latino/Admixed American, East Asian) and adjusted by principal components and percentage of admixture, leading to the generation of 56 PRS models. Logistic regression analysis was employed to predict PD status adjusted either by gender, age, and 10 PCs or by gender, age, and percentage of admixture **(Figure 1)**. This model was standardized to a Z-score with a mean of 0 and a standard deviation of 1. After calculating the allele counts of each variant between cases and controls, we calculated the mean effect of each variant by multiplying the allele count difference by the beta coefficient, or effect size, to estimate the average impact of each variant’s allele count difference on disease phenotype. We focused on variants with the most significant impact for PRS prediction (variants with the highest mean effect). The results were visualized through heatmaps for ancestry comparisons, density plots displaying probabilities of disease, forest plots for magnitude of effects comparison per summary statistics, area under the curve (AUC) and receiver operating characteristic curve (ROC) assessments for sensitivity and specificity. Finally, UpSet visualizations were used to display heterogeneity estimated across known loci and multiple ancestries.

### Model 2: Multi Ancestry Best-Fit PRS approach

#### Ancestry-specific summary statistics generation

For this model we used Kim et al., 2024^6^ summary statistics from the latest multi-ancestry PD GWAS meta-analysis, that includes four populations. The European cohort, was composed of data from Nalls et al., 2019^1^, including 1,467,312 individuals; 56,306 cases (including proxy cases), 1,411,006 controls, and a Finnish cohort of 95,683 participants of which 1,587 were cases and 94,096 controls. The East Asian population combined data from Foo et al., 2020^2^, and *23andMe*, totaling 183,802 individuals of which 7,046 were cases and 176,756 controls. The Latino/Admixed American data was generated merging Loesch et al., 2021^3^ and *23andMe*, encompassing 584,660 individuals, of which 582,220 were controls and 2,440 cases. Additionally, the African admixed population, which derived solely from *23andMe*, consisted of 194,273 individuals, including 288 cases and 193,985 controls. A comprehensive analysis was conducted which aggregated summary statistics across all studies, including a total of 2,525,730 individuals, of which 49,049 were PD cases, 18,618 proxy cases and 2,458,063 controls, highlighting the substantial scope and diversity of the data integrated into this meta-analysis.

#### Multi-ancestry best-fit polygenic risk score calculation

PRS were computed using PRSice-2^32^. We implemented a multi-step process to estimate the cumulative genetic risk attributed to a set of SNPs based on p-value thresholding for each cohort by using multi-ancestry GWAS summary statistics by Kim et al., 2024^6^ (**Figure 1**). PRSice-2 was used to select independent genetic variants following default clumping settings. This approach includes adhering to standardized values (250 kb window, population-specific LD estimation, and an LD threshold of r² < 0.1). The selection was rigorously tailored to each ancestry, facilitating an ancestry-specific assessment.

Subsequently, we implemented a multi p-value thresholding approach to determine the most informative SNPs for inclusion in the PRS, ranging from inclusive (P < 0.5) to stringent (P < 1e-8) criteria. This facilitated the evaluation of PRS predictive performance at varying levels of SNP inclusion. For each cohort, the PRS was calculated by summing the alleles associated with PD and weighted by the effect sizes reported by Kim et al., 2024^6^.

For each cohort we calculated the best p-value threshold for SNP inclusion versus PD risk. We adjusted the model by a disease prevalence of 0.005, sex, age, and PCs. Results were compared between cohorts summarizing the best-fit model for each ancestry including its thresholds and number of included SNPs. We computed Odds ratios (OR) along with their corresponding 95% confidence intervals (CI) for each ancestry group. We visually depicted the performance of the models through both bar plots and heatmaps.

### Sample size and power calculations

We conducted power calculations to estimate the sample size needed to achieve 80% power with a significance level of 0.05, using the methodology proposed by Dudbridge et al.^33^ (additional details can be found at https://github.com/DudbridgeLab/avengeme/). These estimations were made considering the 90 risk variants and the heritability estimates reported in Nalls et al. 2019 (defined as the percentage of the phenotype attributed to genetic variation, h^2^ = 22%) at a 1% PD prevalence. The required sample size for PRS to predict disease status was 544 individuals, so we selected cohorts with more than 500 participants. After fulfilling this criteria, we calculated the power that each of them had following the values stated in Nalls et al. 2019^1^; a Pseudo R2 of 0.054, and an OR of 2.03. (see **Table 1** and https://github.com/GP2code/multiancestry-PRS_PRSice; doi:10.5281/zenodo.11110944).

## RESULTS

### Risk estimates show expected high levels of heterogeneity in predicting disease status across diverse ancestry populations

In analyzing the distribution patterns of the 90 risk alleles from Nalls et al. 2019^1^ across the seven ancestry cohorts under study, significant heterogeneity was observed among these predictors following standardization of the effect allele for each estimate. Differences between ancestries were evident, including the number of valid predictors **(Supplementary Table 2)**, directionality, frequency, and magnitude of effect **(Figure 2, Supplementary Table 3)**.

**Figure 2:**
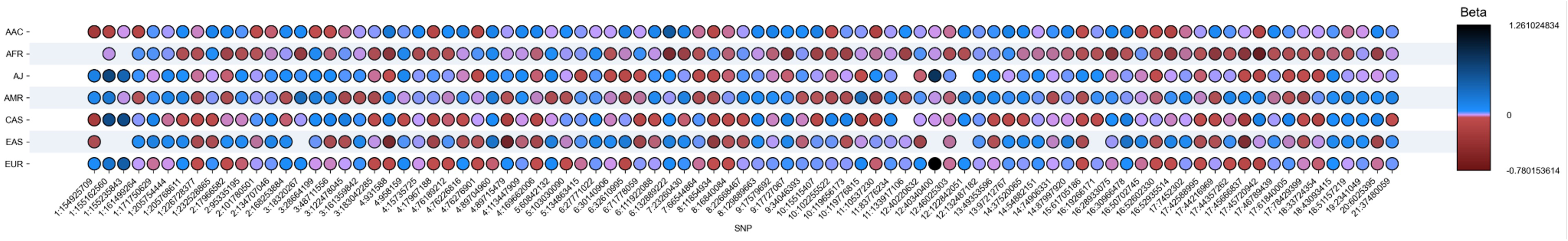
Upset plot showing risk heterogeneity across multiple ancestries. The 90 risk variants are represented in this plot in a granular way. The Y axis represents each ancestry populations and the X axis the 90 risk variants. The color bar shows the magnitude of effects as log of the odd ratio (beta value) and directionality, with red color denoting negative directionality, and purple and blue colors denoting positive directionality.

Further analysis was conducted to determine the individual effect size contributions of genetic variants within each population. These analyses, detailed in **Supplementary Table 4,** uncovered discrepancies among the 90 variants not only in effect size, as depicted in **Figure 2**, but also in the primary variants influencing PRS across populations. In terms of the largest effect contribution, *LRRK2* G2019S and *GBA1* N370S exhibit the most substantial effects in European and Ashkenazi Jewish populations, with the effect in Ashkenazi Jewish being considerably higher. Conversely, *SNCA* (rs356182) emerges as the principal variant for East Asian, Central Asian, African, and African admixed populations. Notably, in Latino/Admixed American populations, *LRRK2* G2019S is the foremost contributor followed by *SNCA* (rs356182), presenting a different pattern than the one observed in the other cohorts.

Regarding the overall number of variants, the East Asian population showed the fewest number of valid predictors **(Figure 2, Supplementary Table 2)**, with 84 imputed variants out of 90, followed by African (88), Ashkenazi Jewish (88), and Central Asian populations (89). European, African admixed, and Latino/Admixed American populations each displayed 90 valid imputed predictors. This result serves as a proof of concept, suggesting the existence of varying linkage disequilibrium risk patterns associated with PD across diverse populations. It underscores the significant amount of genetic variability that remains unexplored in understanding disease risk.

### Conventional polygenic risk scores performance across diverse ancestries

European GWAS-derived PRS models adjusted by sex, age and PCs and including the 90 risk predictors from Nalls et al. 2019 ^1^ exhibited variable predictive accuracy across all ancestries. In European ancestry populations (positive control), this model achieved an OR of 1.60 (95% CI: 1.54-1.70, p < 0.0001) **(Table 2a)**, with an AUC of 0.63, confirming the expected predictability ^1^. The Ashkenazi Jewish population exhibited the highest OR of 1.96 (95% CI: 1.69-2.25, p < 0.0001) **(Table 2a)**, accompanied by an AUC of 0.68 **(Table 4)**, suggesting a comparable predictive capability within this group generally enriched with *LRRK2* G2019S and *GBA1* N370S carriers which are major contributors to the PRS. A similar predictive outcome extended to other ancestries, including East Asians with an OR of 1.37 (95% CI: 1.32-1.42, p < 0.0001) **(Table 2a)** and AUC of 0.62 **(Table 4)**, despite having the lowest number of valid predictors (84) within the cohorts studied. The PRS model for African ancestries yielded an OR of 1.38 (95% CI: 1.12-1.63) **(Table 2a)** with an AUC of 0.54, low sensitivity (0.09) and high specificity **(Table 4)**, pointing to a substantial imbalance in predictive performance. The African Admixed, Latino/Admixed American and Central Asian populations achieved a statistically significant association (p-values of < 0.0001) with ORs of 1.57 (95% CI:1.29 - 1.91), 1.77 (95% CI:1.36 - 2.30), and 1.72 (95% CI:1.32 - 2.30) **(Table 2a)** respectively, but without adequate discriminative abilities as shown by the ROC curve associated estimates seen in **Table 4 and Supplementary Figures 3a-b).**

**Table 2.a.** Polygenic risk scores vs. PD status adjusted by age, sex, and PCs across multiple ancestry populations.

| Ancestry | Summary statistics | Odds Ratio (95% CI) | p value |
| --- | --- | --- | --- |
| EUR | AAC | 1.15 (1.11- 1.19) | <0.0001 |
|  | EUR | 1.60 (1.54 - 1.70) | <0.0001 |
|  | AMR | 1.27 (1.23 - 1.32) | <0.0001 |
|  | EAS | 1.37 (1.32 - 1.42) | <0.0001 |
| AAC | AAC | 1.37 (1.14 - 1.70) | 0.001 |
|  | EUR | 1.57 (1.29 - 1.91) | <0.0001 |
|  | AMR | 1.15 (0.96 - 1.39) | 0.120 |
|  | EAS | 1.36 (1.13 - 1.65) | 0.010 |
| AMR | AAC | 1.52 (1.16 - 1.99) | 0.002 |
|  | EUR | 1.77 (1.36 - 2.30) | <0.0001 |
|  | AMR | 1.37 (1.06 - 1.75) | 0.016 |
|  | EAS | 1.678 (1.29 - 2.19) | <0.0001 |
| EAS | AAC | 1.11 (1.01 - 1.23) | 0.042 |
|  | EUR | 1.50 (1.35 - 1.67) | <0.0001 |
|  | AMR | 1.21 (1.09 - 1.37) | <0.0001 |
|  | EAS | 1.30 (1.17 - 1.44) | <0.0001 |
| AFR | AAC | 1.26 (1.10 - 1.50) | 0.009 |
|  | EUR | 1.38 (1.12 - 1.63) | 0.001 |
|  | AMR | 1.15 (0.97 - 1.36) | 0.121 |
|  | EAS | 1.32 (1.10 - 1.57) | 0.002 |
| AJ | AAC | 1.03 (0.91 - 1.18) | 0.629 |
|  | EUR | 1.96 (1.69 - 2.25) | <0.0001 |
|  | AMR | 1.24 (1.08 - 1.41) | 0.002 |
|  | EAS | 1.30 (1.13 - 1.48) | <0.0001 |
| CAS | AAC | 1.20 (0.911- 1.58) | 0.194 |
|  | EUR | 1.72 (1.32 - 2.30) | <0.0001 |
|  | AMR | 1.46 (1.12 - 1.90) | 0.005 |
|  | EAS | 1.27 (0.99 - 1.62) | 0.067 |

Across all ancestries studied, PRS developed from ancestry-specific summary statistics based on the 90 risk predictors did not outperform European-based PRS models using Nalls et al., 2019^1^ summary statistics, most likely due to limited statistically powered population-specific summary statistics and population differences in LD risk patterns which may not be representative of our current understanding of disease risk based on European populations (**Table 2a-2b, Supplementary Table 2, Figure 3**).

**Figure 3:**
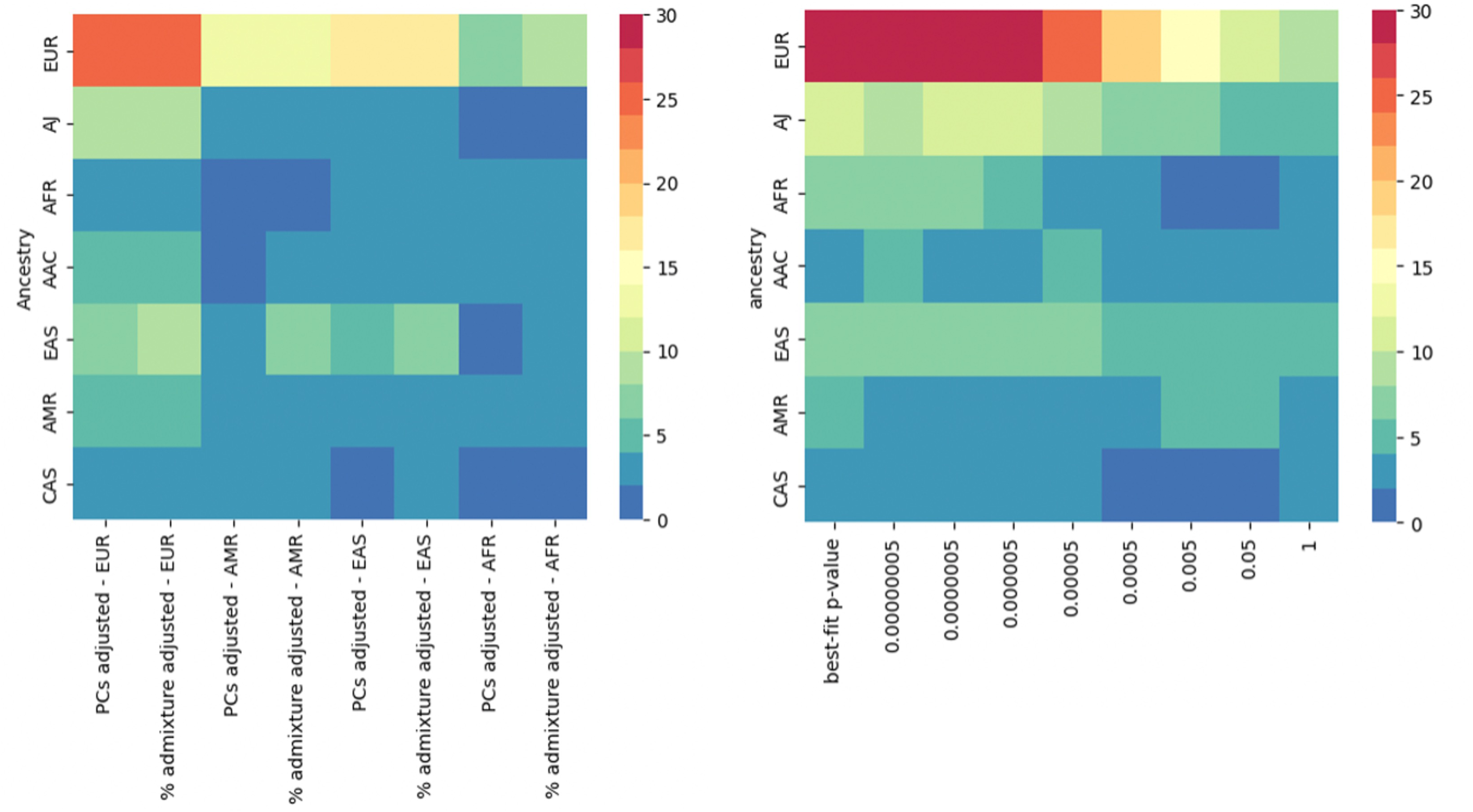
Polygenic risk score performance for predicting disease status in the different models. Left panel shows results for conventional PRS calculation and right panel the best fit PRS model. The Y axis represents individual level data, and the X axis represents the two different PRS approaches per population-specific summary statistics. The color bar indicates the magnitude of effect as zeta value (beta/se). The darker the color, the larger the magnitude of effect. The asterisks indicate statistical significance of P value.

**Table 2.b.**
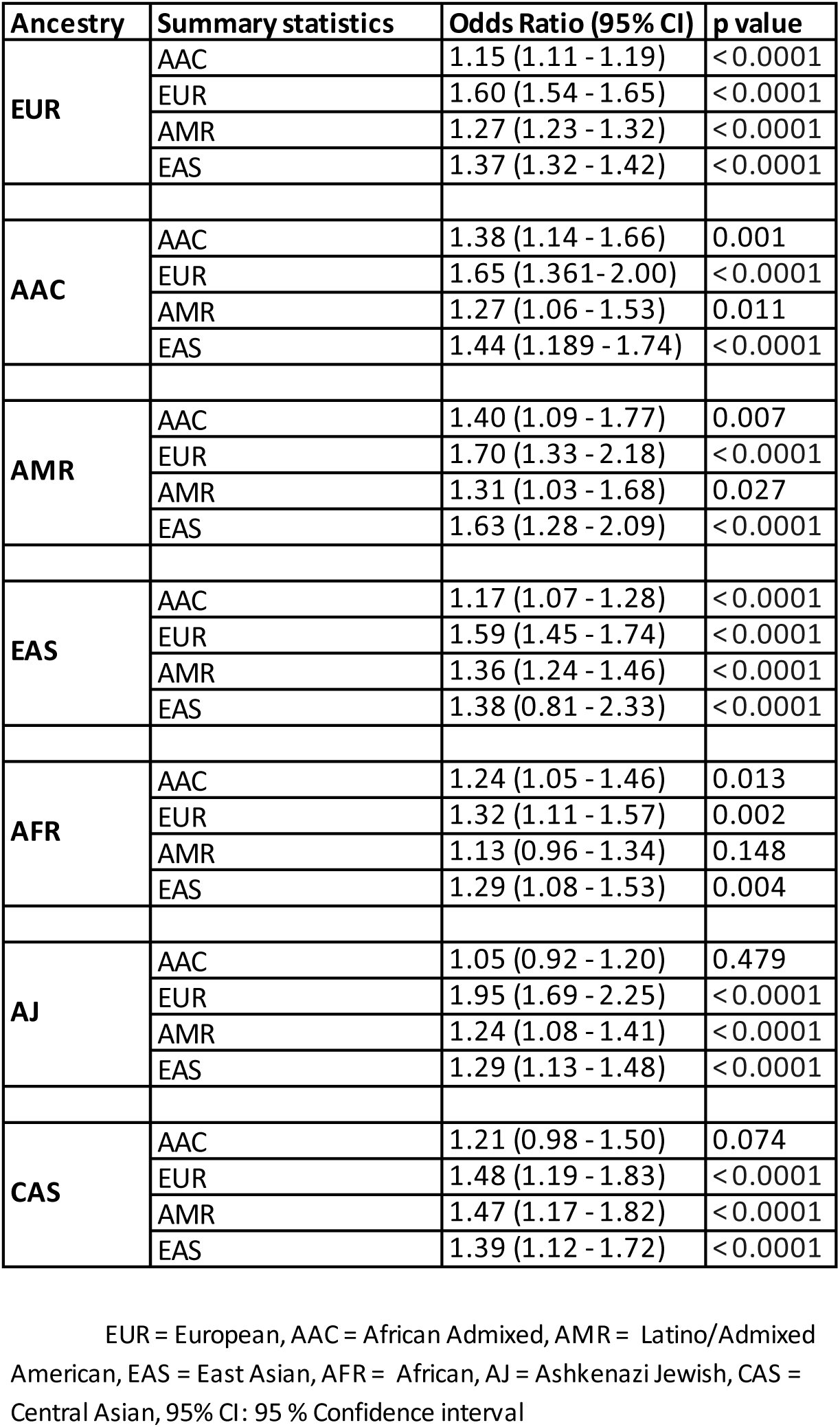
Polygenic risk scores vs. PD status adjusted by age, sex, and percentage of ancestry across multiple ancestry populations.

| Ancestry | Summary statistics | Odds Ratio (95% CI) | p value |
| --- | --- | --- | --- |
| EUR | AAC | 1.15 (1.11 - 1.19) | <0.0001 |
|  | EUR | 1.60 (1.54 - 1.65) | <0.0001 |
|  | AMR | 1.27 (1.23 - 1.32) | <0.0001 |
|  | EAS | 1.37 (1.32 - 1.42) | <0.0001 |
| AAC | AAC | 1.38 (1.14 - 1.66) | 0.001 |
|  | EUR | 1.65 (1.361- 2.00) | <0.0001 |
|  | AMR | 1.27 (1.06 - 1.53) | 0.011 |
|  | EAS | 1.44 (1.189 - 1.74) | <0.0001 |
| AMR | AAC | 1.40 (1.09 - 1.77) | 0.007 |
|  | EUR | 1.70 (1.33 - 2.18) | <0.0001 |
|  | AMR | 1.31 (1.03 - 1.68) | 0.027 |
|  | EAS | 1.63 (1.28 - 2.09) | <0.0001 |
| EAS | AAC | 1.17 (1.07 - 1.28) | <0.0001 |
|  | EUR | 1.59 (1.45 - 1.74) | <0.0001 |
|  | AMR | 1.36 (1.24 - 1.46) | <0.0001 |
|  | EAS | 1.38 (0.81 - 2.33) | <0.0001 |
| AFR | AAC | 1.24 (1.05 - 1.46) | 0.013 |
|  | EUR | 1.32 (1.11 - 1.57) | 0.002 |
|  | AMR | 1.13 (0.96 - 1.34) | 0.148 |
|  | EAS | 1.29 (1.08 - 1.53) | 0.004 |
| AJ | AAC | 1.05 (0.92 - 1.20) | 0.479 |
|  | EUR | 1.95 (1.69 - 2.25) | <0.0001 |
|  | AMR | 1.24 (1.08 - 1.41) | <0.0001 |
|  | EAS | 1.29 (1.13 - 1.48) | <0.0001 |
| CAS | AAC | 1.21 (0.98 - 1.50) | 0.074 |
|  | EUR | 1.48 (1.19 - 1.83) | <0.0001 |
|  | AMR | 1.47 (1.17 - 1.82) | <0.0001 |
|  | EAS | 1.39 (1.12 - 1.72) | <0.0001 |

### Adjustment for percentage of admixture versus principal components does not generally ameliorate polygenic risk score performance

We aimed to further adjust by the potential variability caused by admixture patterns. As demonstrated by the results highlighted in **Table 2b** and schematically depicted in **Figure 3**, PRS performance for each ancestry does not improve when the models are adjusted by the percentage of admixture versus PCs, except in the case of the East Asian population, were the model adjusted by percentage of admixture displayed an OR of 1.38 (0.81-2.33, p< 0.0001) compared to PCs adjustment. This observation suggests that adjustment by PCs sufficiently accounts for ancestry in the conventional PRS model and for most of the assessed populations.

### Multi-ancestry best-fit polygenic risk score models surpass conventional approaches except in admixed populations

Our multi-ancestry best-fit PRS model based on p-value thresholding demonstrates varied effectiveness across ancestries, enhancing genetic risk prediction particularly in European, Ashkenazi Jewish, and East Asians, and showing a substantial improvement for African populations **(Table 3**, **Figure 3b, Supplementary Figure 5**). European populations achieved a high level of predictive accuracy with a p-value threshold of 5E-07, and 266 selected SNPs to reach an OR of 1.66 (1.60 - 1.71) **(Table 3)**. Similarly to the conventional PRS, Ashkenazi Jewish displayed stronger association with an OR of 2.81 (2.33 - 3.39), utilizing 459 SNPs and a threshold of 5E-06 **(Table 3)**. The East Asian population showed the highest efficiency with a low threshold of 5E-08, using the lower number of SNPs (211 SNPs) and achieving an OR of 1.47 (1.33 - 1.62) **(Table 3)**. However, for African Admixed, Latino/Admixed American, and Central Asian populations the multi-ancestry best-fit PRS model performed worse than conventional PRS. No major differences in AUC estimates were observed when comparing Multi-ancestry best-fit PRS with the conventional model. Figure 4 compares ROC curves across ancestries.

**Figure 4:**
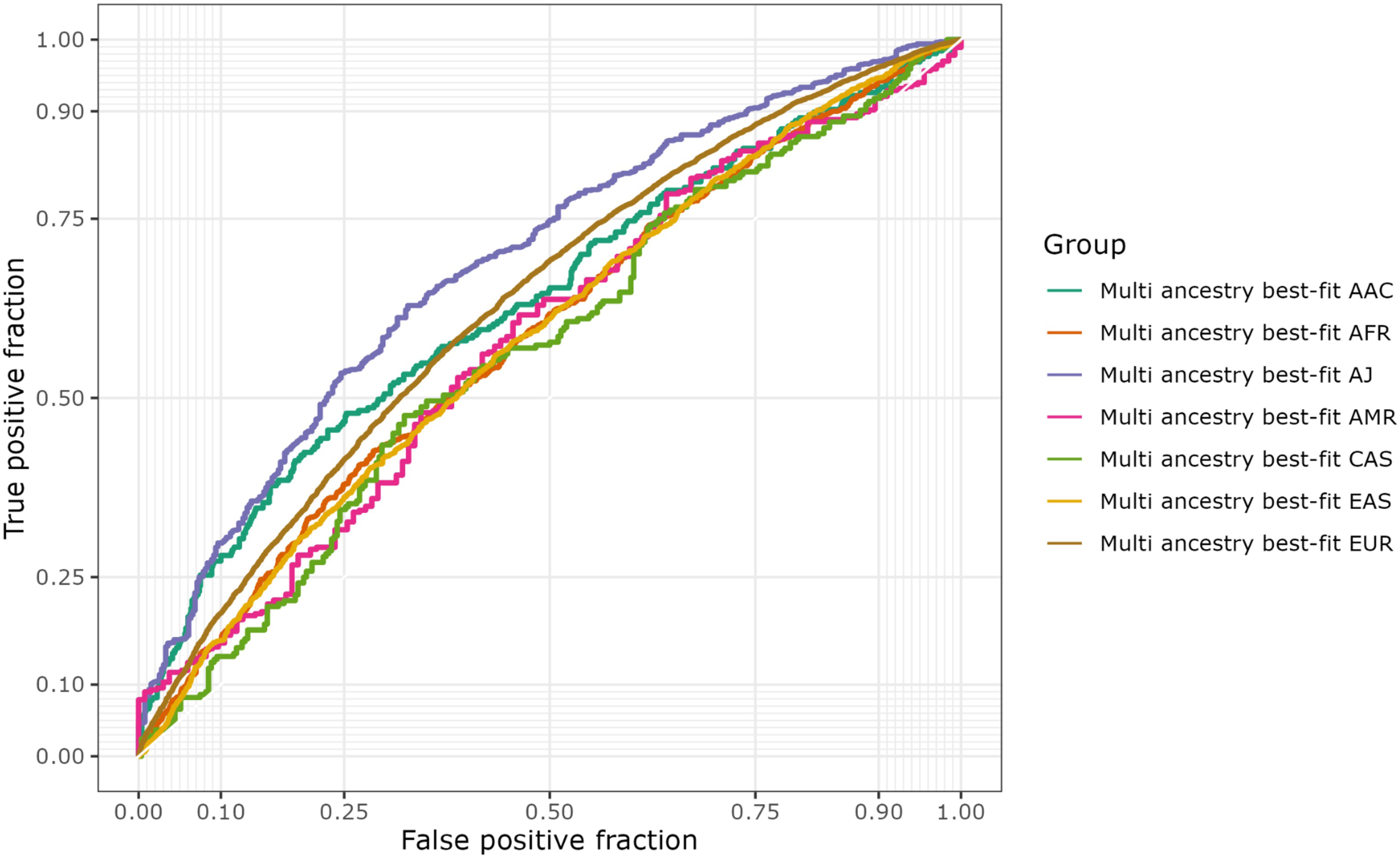
Polygenic risk score model performance evaluation for Multi-ancestry best fit PRS. The ROC curve depicts an evaluation of the PRS best fit model’s performance for each target data population, represented in a different colored curve. The true positive rate is plotted on the Y axis against the false positive rate on the X axis. The sensitivity of the model increases with increasing Y value. The specificity (1-specificity) of the model decreases as the X value increases.

**Table 3.**
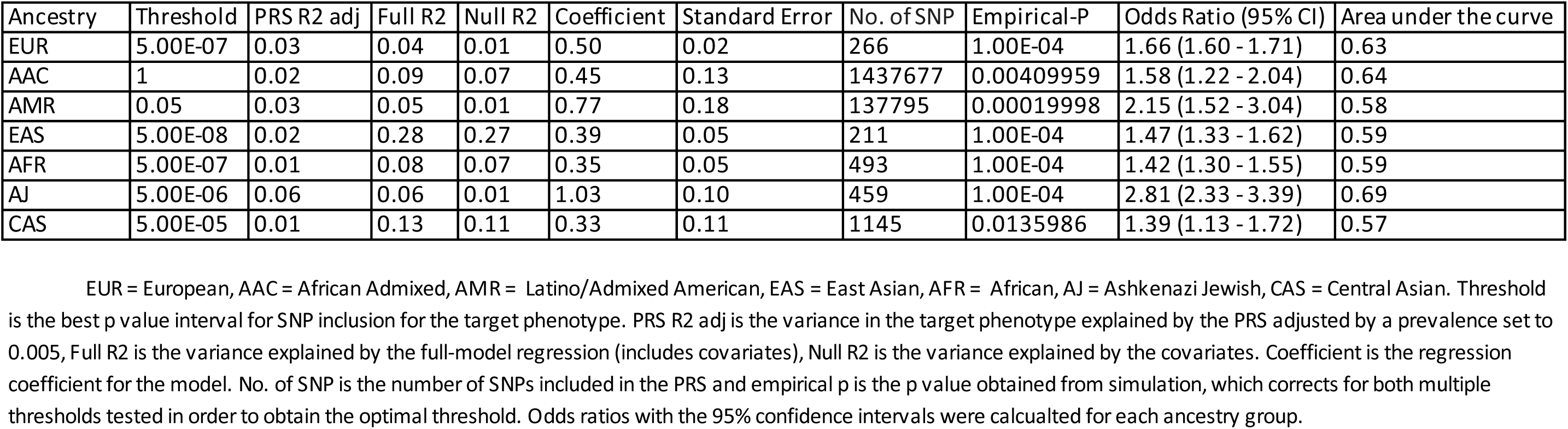
Multi-ancestry best-fit polygenic risk score analysis.

| Ancestry | Threshold | PRS R2 adj | Full R2 | Null R2 | Coefficient | Standard Error | No. of SNP | Empirical-P | Odds Ratio (95% CI) | Area under the curve |
| --- | --- | --- | --- | --- | --- | --- | --- | --- | --- | --- |
| EUR | 5.00E-07 | 0.03 | 0.04 | 0.01 | 0.50 | 0.02 | 266 | 1.00E-04 | 1.66 (1.60 - 1.71) | 0.63 |
| AAC | 1 | 0.02 | 0.09 | 0.07 | 0.45 | 0.13 | 1437677 | 0.00409959 | 1.58 (1.22 - 2.04) | 0.64 |
| AMR | 0.05 | 0.03 | 0.05 | 0.01 | 0.77 | 0.18 | 137795 | 0.00019998 | 2.15 (1.52 - 3.04) | 0.58 |
| EAS | 5.00E-08 | 0.02 | 0.28 | 0.27 | 0.39 | 0.05 | 211 | 1.00E-04 | 1.47 (1.33 - 1.62) | 0.59 |
| AFR | 5.00E-07 | 0.01 | 0.08 | 0.07 | 0.35 | 0.05 | 493 | 1.00E-04 | 1.42 (1.30 - 1.55) | 0.59 |
| AJ | 5.00E-06 | 0.06 | 0.06 | 0.01 | 1.03 | 0.10 | 459 | 1.00E-04 | 2.81 (2.33 - 3.39) | 0.69 |
| CAS | 5.00E-05 | 0.01 | 0.13 | 0.11 | 0.33 | 0.11 | 1145 | 0.0135986 | 1.39 (1.13 - 1.72) | 0.57 |

**Table 4:** Area under the curve calculation across multiple ancestry population.

| Ancestry | Summary statistics | Area under the curve | Accuracy | 95% CI Accuracy | Balanced Accuracy | Sensitivity | Specificity |
| --- | --- | --- | --- | --- | --- | --- | --- |
| EUR | AAC | 0.54 | 0.61 | (0.60-0.62) | 0.50 | 1.00 | 0.00 |
|  | EUR | 0.63 | 0.63 | (0.63 - 0.64) | 0.56 | 0.88 | 0.24 |
|  | AMR | 0.57 | 0.61 | (0.60-0.62) | 0.51 | 0.97 | 0.05 |
|  | EAS | 0.59 | 0.61 | (0.61 - 0.62) | 0.52 | 0.95 | 0.09 |
| AAC | AAC | 0.59 | 0.76 | (0.73 - 0.78) | 0.50 | 0.00 | 1.00 |
|  | EUR | 0.63 | 0.76 | (0.73 - 0.78) | 0.50 | 0.01 | 0.99 |
|  | AMR | 0.56 | 0.76 | (0.73 - 0.78) | 0.50 | 0.00 | 1.00 |
|  | EAS | 0.61 | 0.76 | (0.73 - 0.78) | 0.50 | 0.00 | 1.00 |
| AMR | AAC | 0.57 | 0.73 | (0.69-0.76) | 0.50 | 1.00 | 0.00 |
|  | EUR | 0.62 | 0.72 | (0.68 - 0.76) | 0.51 | 0.99 | 0.02 |
|  | AMR | 0.57 | 0.73 | (0.68 - 0.76) | 0.50 | 1.00 | 0.00 |
|  | EAS | 0.61 | 0.73 | (0.68 - 0.76) | 0.50 | 1.00 | 0.00 |
| EAS | AAC | 0.57 | 0.60 | (0.59 – 0.62) | 0.52 | 0.07 | 0.96 |
|  | EUR | 0.62 | 0.62 | (0.61 – 0.64) | 0.56 | 0.25 | 0.87 |
|  | AMR | 0.58 | 0.60 | (0.59 - 0.62) | 0.50 | 0.07 | 0.95 |
|  | EAS | 0.56 | 0.60 | (0.58 – 0.61) | 0.50 | 0.02 | 0.98 |
| AFR | AAC | 0.54 | 0.65 | (0.63 - 0.66) | 0.50 | 0.00 | 1.00 |
|  | EUR | 0.54 | 0.65 | (0.63 - 0.66) | 0.50 | 0.00 | 1.00 |
|  | AMR | 0.51 | 0.65 | (0.63 - 0.66) | 0.50 | 0.00 | 1.00 |
|  | EAS | 0.53 | 0.65 | (0.63 - 0.66) | 0.50 | 0.00 | 1.00 |
| AJ | AAC | 0.54 | 0.69 | (0.66-0.71) | 0.50 | 1.00 | 0.00 |
|  | EUR | 0.68 | 0.70 | (0.67 - 0.72) | 0.54 | 0.94 | 0.15 |
|  | AMR | 0.56 | 0.69 | (0.66 - 0.71) | 0.50 | 1.00 | 0.00 |
|  | EAS | 0.57 | 0.69 | (0.66 - 0.71) | 0.50 | 1.00 | 0.00 |
| CAS | AAC | 0.55 | 0.53 | (0.49 - 0.57) | 0.53 | 0.47 | 0.60 |
|  | EUR | 0.56 | 0.53 | (0.49 - 0.57) | 0.53 | 0.49 | 0.57 |
|  | AMR | 0.58 | 0.56 | (0.52 - 0.60) | 0.56 | 0.53 | 0.59 |
|  | EAS | 0.56 | 0.54 | (0.50-0.58) | 0.54 | 0.51 | 0.57 |

## DISCUSSION

To our knowledge, this study represents the first comprehensive assessment of PRS in predicting PD risk in a multi-ancestry context. While previous genetics research has primarily focused on studying populations of European ancestry^1,34,35^, our study expands on previous knowledge by comparing the performance of conventional PRS across seven ancestry populations while implementing a novel and refined multi-ancestry best-fit polygenic risk score approach to enhance prediction applicability in a global setting.

Our study reveals that although our understanding of PD risk is predominantly derived from European genetic studies, the conventional PRS model utilizing summary statistics from this population shows to some extent applicability across diverse populations, including Ashkenazi Jewish (harboring certain levels of European ancestry and enriched with *LRRK2* and *GBA1* carriers) and East Asians. Interestingly, adjusting the model to account for percentage of admixture versus conventional PCs does not significantly improve its predictive accuracy. Furthermore, PRS models derived from the 90 risk predictors originating from European populations and constructed using estimates from population-specific summary statistics failed to enhance predictions. This is likely attributed to the scarcity of statistically robust population-specific summary statistics and variations in LD risk patterns among populations, potentially diverging from the prevailing understanding of disease risk as established in European populations^36^.

To reconcile these discrepancies and enhance our ability to forecast risk, we devised a best-fit multi-ancestry PRS approach based on p-value thresholding by leveraging multi-ancestry GWAS data as the base to select the best set of cumulative SNPs discriminating cases from controls. Our approach optimizes the model notably for less admixed populations. Taking into consideration p-value thresholding and population-specific LD patterns, we managed to enhance the model’s precision in the context of risk yet seems less effective in more genetically admixed populations. The varied performance of the best-fit PRS across different ancestries exemplifies the challenge that a ‘one size fits all’ approach presents in genetic research, advocating for a more nuanced strategy in precision medicine that accommodates the rich genetic variability of global populations.

The results observed in East Asian aligns with the work of Foo et al., 2020 ^2^ and supports the cross population applicability of PRS, that has already been evidenced in this population in the context of Alzheimer’s disease^37^, breast cancer^38^, and colorectal cancer^39^. The major contributor for the PRS in this cohort was *SNCA* (rs356182), with an absolute mean effect twice as high as *LRRK2* G2019S, the highest SNP in Europeans (**Supplementary Table 4**). This result is particularly compelling as Europeans and East Asian genetics ancestries are very different -illustrated in ancestry prediction models- (**Supplementary Figure 1**), and contrasts with the hypothesis that the accuracy of PRS depends on genetic ancestry relatedness ^13^.

Several limitations should be acknowledged. Due to limited information on heritability, disease prevalence, and risk predictors for non-European ancestries, sample size power calculations were performed using current estimates from the European population as a reference. Consequently, this may result in a biased estimate regarding the sample size required to predict disease status across diverse ancestries. Additionally, the estimates of our models are influenced by the number of available SNPs in each dataset, which introduces bias. This bias arises from variations in the quality and completeness of SNP imputation across different populations, where some of them may have a larger number of imputed variants e.g., 90 for Latino/Admixed American compared to others (e.g., 84 for East Asian and 88 for African). This is due to differences in variant frequencies in which common risk variation contributing to disease in Europeans is rare when assessed in other populations and therefore impacted by limited imputation. An additional important limitation is the absence of individual-level replication datasets per ancestry. The lack of replication data hampers the robustness and generalizability of our findings across different individual level data from diverse ancestral populations. Furthermore, the scalability of our framework is hindered by the absence of accurate and well-powered ancestry-specific summary statistics for each population in our study, like the African Admixed summary statistics that were used in this study.

To overcome these limitations future research should prioritize larger sample sizes for individual-level datasets per ancestry, and availability of well-powered ancestry-specific summary statistics. Moreover, new strategies in PRS construction, like incorporating local ancestry estimates^40^, could significantly improve outcomes in highly admixed populations. This approach enables us to utilize summary statistics from the ancestry PRS panel corresponding to the specific chromosomal region of the individual under risk inference, mitigating inflation or deflation caused by ancestry-specific risk alleles. Studying biomarker-defined PD cohorts, as opposed to those diagnosed based on clinical diagnostic criteria, is crucial, as at least 5% of individuals diagnosed with PD do not demonstrate neuronal alpha-synuclein, which is required for definitive diagnosis^41^. Additionally, applying multi-modality machine learning (ML) approaches^12^ that combine adjusted transcriptomics, genetics, and clinical data into a predictive model, could provide a more comprehensive understanding of PD risk and improve prediction accuracy across diverse ancestries. By utilizing ML algorithms such as deep learning, complex patterns and interactions that may not be evident when using individual data modalities alone can have the potential to enhance the precision and applicability of PD risk assessment models. This would lead to improved risk prediction and personalized strategies for prevention, diagnosis, and treatment for all.

This study is the first to comprehensively compare two PRS approaches to predict PD risk across seven ancestries, comparing the conventional model based on genetic risk defined through GWAS conducted in European populations versus a more refined multi-ancestry best-fit approach. Despite these efforts, our study reveals the need for additional data and novel approaches, such as the inclusion of local ancestry information to improve PRS applicability in highly admixed groups. Here, we confirm the transferability of European-derived PRS models to other ancestries, opening avenues for their broader use. Integrating clinical and genetic data^9^ with cutting-edge multi-modality machine learning techniques^12^ could reveal complex disease patterns previously unnoticed. Future research, employing composite PRS analysis for optimized SNP selection across ancestries, holds promise for more accurate risk predictions. Such progress is pivotal for advancing PRS precision and its application in PD, potentially revolutionizing prevention, diagnosis, and treatment on a personalized level.

## Data Availability

Data were obtained from the Global Parkinson's Genetics Program (GP2) and is accessible through a partnership with the Accelerating Medicines Partnership in Parkinson's Disease (AMP-PD) and can be requested via the website's application process (https://www.amp-pd.org/). GWAS summary statistics from GP2's release 6 are available for all datasets (doi: 10.5281/zenodo.10472143, https://doi.org/10.5281/zenodo.10472143). 23andMe summary statistics is available upon application through their website (https://research.23andme.com/dataset-access/). GenoTools (version 10; https://github.com/GP2code/GenoTools) was used for genotyping, imputation, quality control, ancestry prediction, and data processing. A secured workspace on the Terra platform was created to conduct genetic analyses using GP2 release 6 data and summary statistics (https://app.terra.bio/). Additionally, all scripts used for this study can be found in the public domain on GitHub (https://github.com/GP2code/multiancestry-PRS_PRSice; doi:10.5281/zenodo.11110944).

doi:10.5281/zenodo.11110944

## Data and Code Availability

Data was obtained from the Global Parkinson’s Genetics Program (GP2) and is accessible through a partnership with the Accelerating Medicines Partnership in Parkinson’s Disease (AMP-PD) and can be requested via the website’s application process (https://www.amp-pd.org/). GWAS summary statistics from GP2’s release 6 are available for all datasets (doi: 10.5281/zenodo.10472143, https://doi.org/10.5281/zenodo.10472143). 23andMe summary statistics is available upon application through their website (https://research.23andme.com/dataset-access/). GenoTools (version 10; https://github.com/GP2code/GenoTools) was used for genotyping, imputation, quality control, ancestry prediction, and data processing. A secured workspace on the Terra platform was created to conduct genetic analyses using GP2 release 6 data and summary statistics (https://app.terra.bio/). Additionally, all scripts used for this study can be found in the public domain on GitHub (https://github.com/GP2code/multiancestry-PRS_PRSice; doi:10.5281/zenodo.11110944).

## Acknowledgements

This work was carried out with the support and guidance of the ‘GP2 Trainee Network’ which is part of the Global Parkinson’s Genetics Program and funded by the Aligning Science Across Parkinson’s (ASAP) initiative. Data used in the preparation of this article were obtained from Global Parkinson’s Genetics Program (GP2). GP2 is funded by the Aligning Science Across Parkinson’s (ASAP) initiative and implemented by The Michael J. Fox Foundation for Parkinson’s Research (https://gp2.org). For a complete list of GP2 members see https://gp2.org.

This research was supported in part by the Intramural Research Program of the NIH, National Institute on Aging (NIA), National Institutes of Health, Department of Health, and Human Services; project number ZIAAG000534, as well as the National Institute of Neurological Disorders and Stroke. This work utilized the computational resources of the NIH HPC Biowulf cluster. (http://hpc.nih.gov)

We are grateful to the Banner Sun Health Research Institute Brain and Body Donation Program of Sun City, Arizona for the provision of human biological materials. The Brain and Body Donation Program has been supported by the National Institute of Neurological Disorders and Stroke (U24 NS072026 National Brain and Tissue Resource for Parkinson’s Disease and Related Disorders), the National Institute on Aging (P30 AG19610 and P30AG072980, Arizona Alzheimer’s Disease Center), the Arizona Department of Health Services (contract 211002, Arizona Alzheimer’s Research Center), the Arizona Biomedical Research Commission (contracts 4001, 0011, 05-901 and 1001 to the Arizona Parkinson’s Disease Consortium) and the Michael J. Fox Foundation for Parkinson’s Research.

We would like to thank the research participants and employees of 23andMe for making this work possible. The following members of the 23andMe Research Team contributed to this study: Stella Aslibekyan, Adam Auton, Elizabeth Babalola, Robert K. Bell, Jessica Bielenberg, Jonathan Bowes, Katarzyna Bryc, Ninad S. Chaudhary, Daniella Coker, Sayantan Das, Emily DelloRusso, Sarah L. Elson, Nicholas Eriksson, Teresa Filshtein, Pierre Fontanillas, Will Freyman, Zach Fuller, Chris German, Julie M. Granka, Alejandro Hernandez, Barry Hicks, David A. Hinds, Ethan M. Jewett, Yunxuan Jiang, Katelyn Kukar, Alan Kwong, Yanyu Liang, Keng-Han Lin, Bianca A. Llamas, Matthew H. McIntyre, Steven J. Micheletti, Meghan E. Moreno, Priyanka Nandakumar, Dominique T. Nguyen, Jared O’Connell, Aaron A. Petrakovitz, G. David Poznik, Alexandra Reynoso, Shubham Saini, Morgan Schumacher, Leah Selcer, Anjali J. Shastri, Janie F. Shelton, Jingchunzi Shi, Suyash Shringarpure, Qiaojuan Jane Su, Susana A. Tat, Vinh Tran, Joyce Y. Tung, Xin Wang, Wei Wang, Catherine H. Weldon, Peter Wilton, Corinna D. Wong.

## Author Contributions

SBC, MBM and HL contributed to the conception and design of the study.

PSA, AOS, IE, SBC, MBM, HL, MAN, KH, MJK, KSL, DV, JK, TPL, MTP, SD, ARP, NRL, JNF, WM, LNK,

MR, NO, CB, IFM contributed to the acquisition and analysis of the data.

PSA, AOS, IE, SBC, MBM, HL, PWC, AFSS, IFM contributed to drafting the text and/or preparing the figures.

All the authors contributed to editing and critically reviewing the manuscript.

