## Supplementary figures for "Insights into Ancestral Diversity in Parkinson’s Disease Risk: A Comparative Assessment of Polygenic Risk Scores"

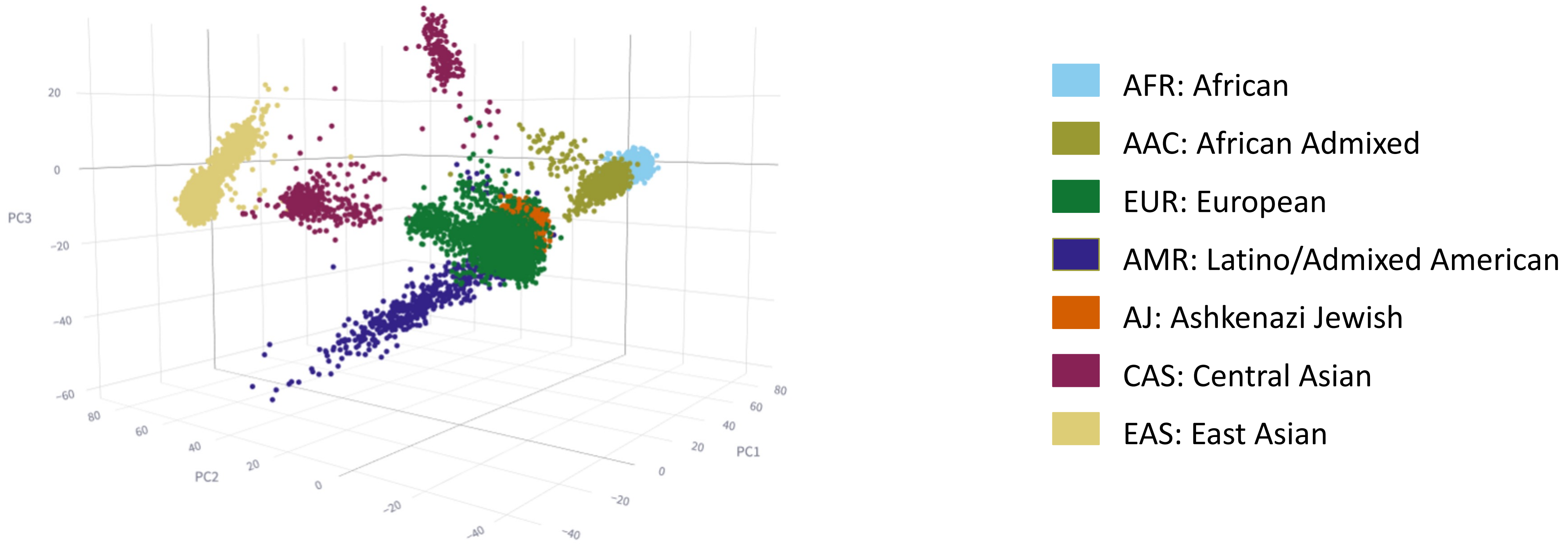


**Supplementary Figure 1: Ancestry prediction model for target data.** Three-dimensional principal components analysis (PCA) was performed to group individuals based on their genetic makeup. Each dot in the figure represents a sample, and the colors depict the ancestral background, as indicated in the color legend.

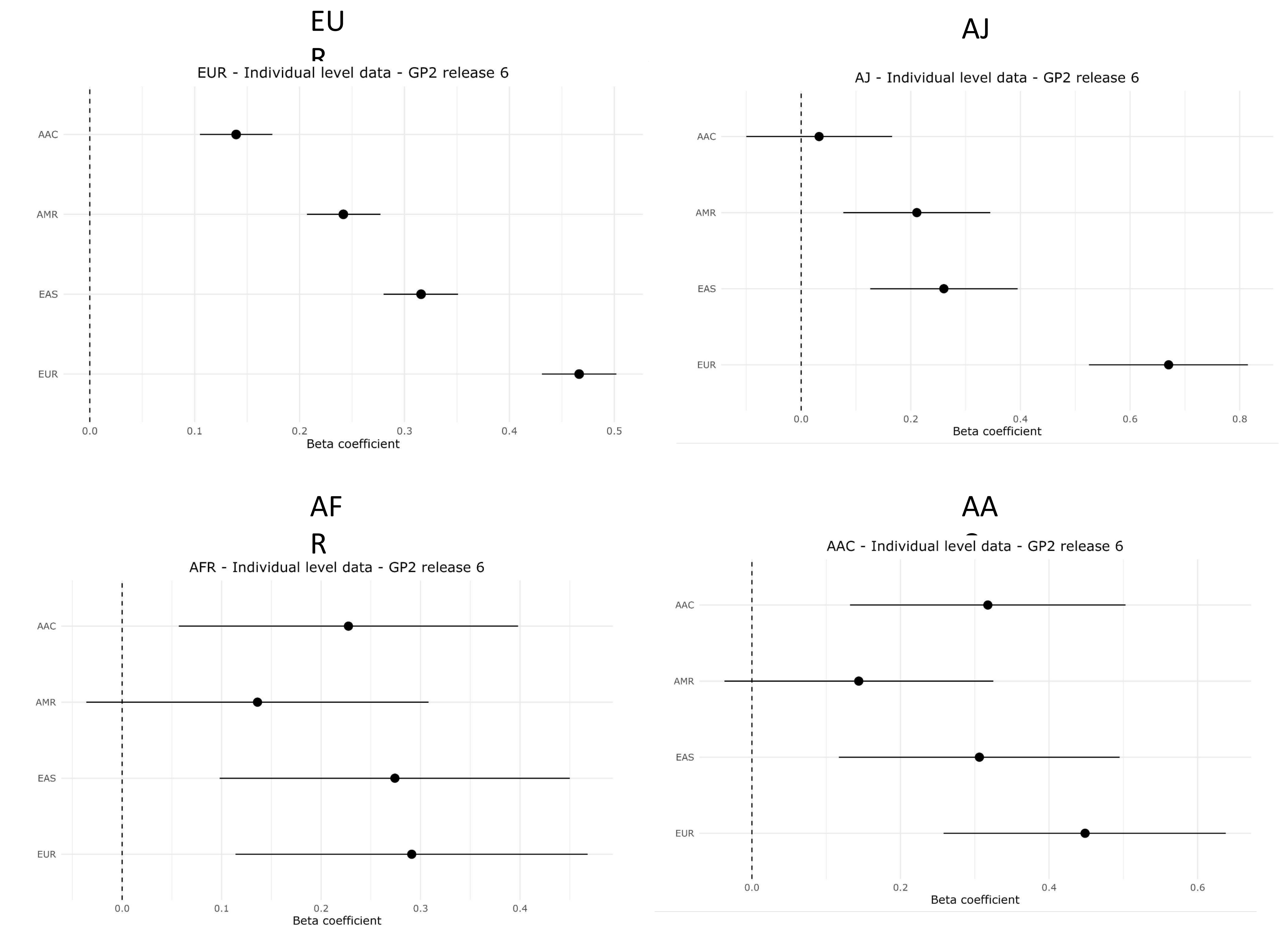


**Supplementary Figure 2a**: **Magnitude of effect across ancestries**
Forest plots comparing the effectiveness of disease prediction across the studied ancestries. Each panel specifically contrasts individual-level data with the population-specific summary statistics: European (EUR), East Asian (EAS), Latino Admixed (AMR), and African Admixed (AAC). On the plots, the x-axis represents the magnitude of effect, the y-axis lists the summary statistics for each group. The dots symbolize the value of the beta coefficient, and horizontal lines depict confidence intervals.

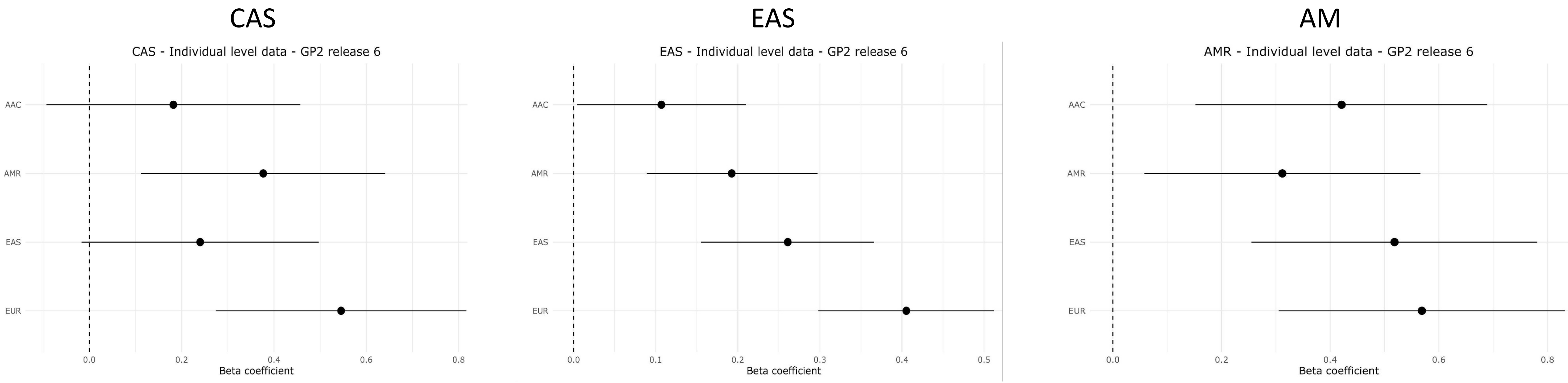


**Supplementary Figure 2b**: **Magnitude of effect across ancestries**
Forest plots comparing the effectiveness of disease prediction across the studied ancestries. Each panel specifically contrasts individual-level data with the population-specific summary statistics: European (EUR), East Asian (EAS), Latino Admixed (AMR), and African Admixed (AAC). On the plots, the x-axis represents the magnitude of effect, the y-axis lists the summary statistics for each group. The dots symbolize the value of the beta coefficient, and the horizontal lines depict confidence intervals.

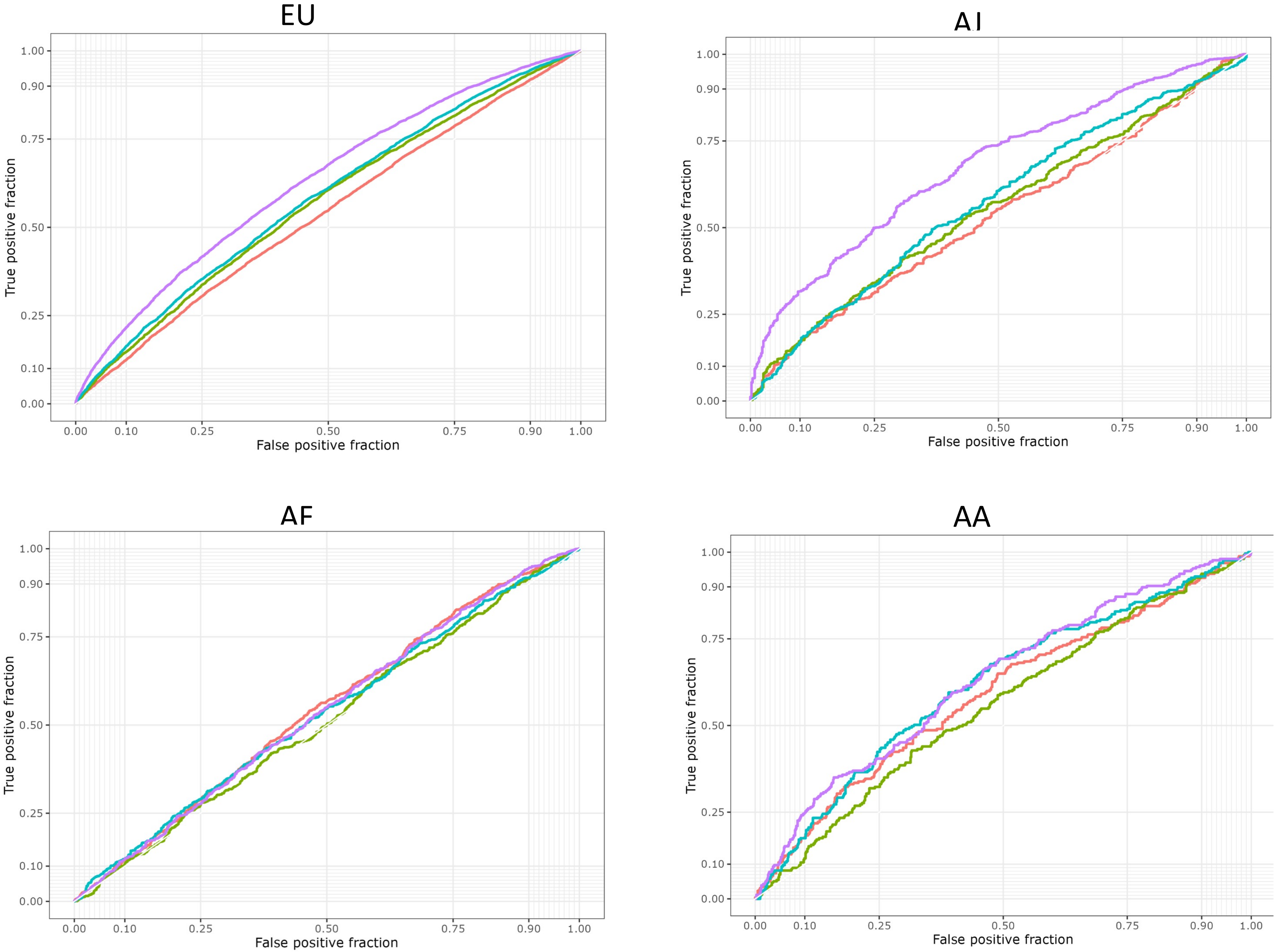


**Supplementary Figure 3a**: **Polygenic risk score model performance evaluation**
The ROC curve depicts an evaluation of the PRS model's performance for each target data population when using the 4 population-specific summary statistics and the multi-ancestry PD summary statistics. Each panel represents a comparison of individual-level data. The true positive rate is plotted on the Y axis against the false positive rate on the X axis. The sensitivity of the model increases with increasing Y value. The specificity (1-specificity) of the model decreases as the X value increases. Each population-specific PRS summary statistics is represented as a curve.

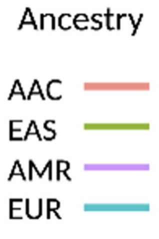

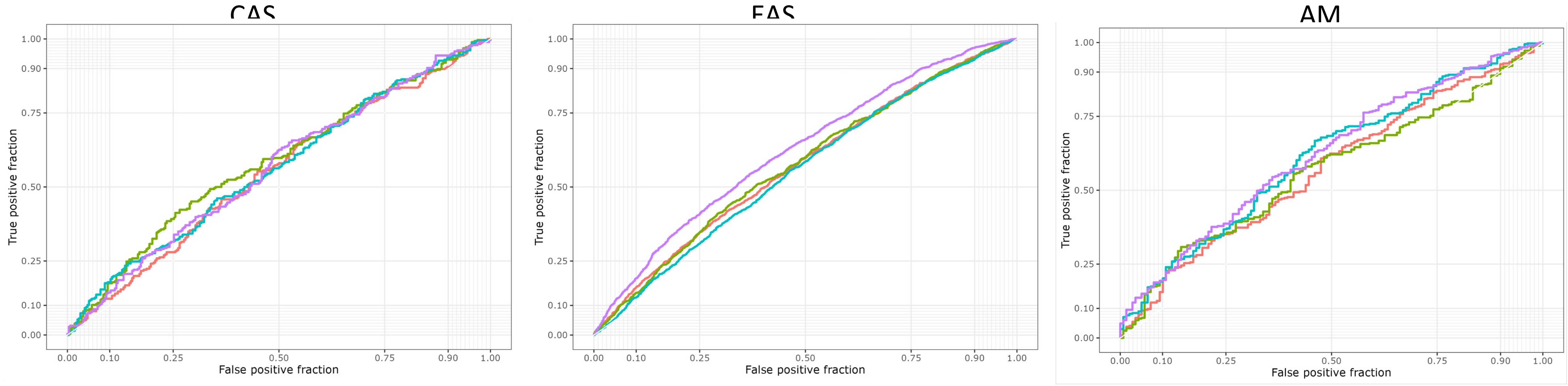


**Supplementary Figure 3b**: Polygenic risk score model performance evaluation

The ROC curve depicts an evaluation of the PRS model's performance for each target data population when using the 4 population-specific summary statistics and the multi-ancestry PD summary statistics. Each panel represents a comparison of individual-level data. The true positive rate is plotted on the Y axis against the false positive rate on the X axis. The sensitivity of the model increases with increasing Y value. The specificity (1-specificity) of the model decreases as the X value increases. Each population-specific PRS summary statistics is represented as a curve.

Ancestry

AAC
EAS
AMR
EUR

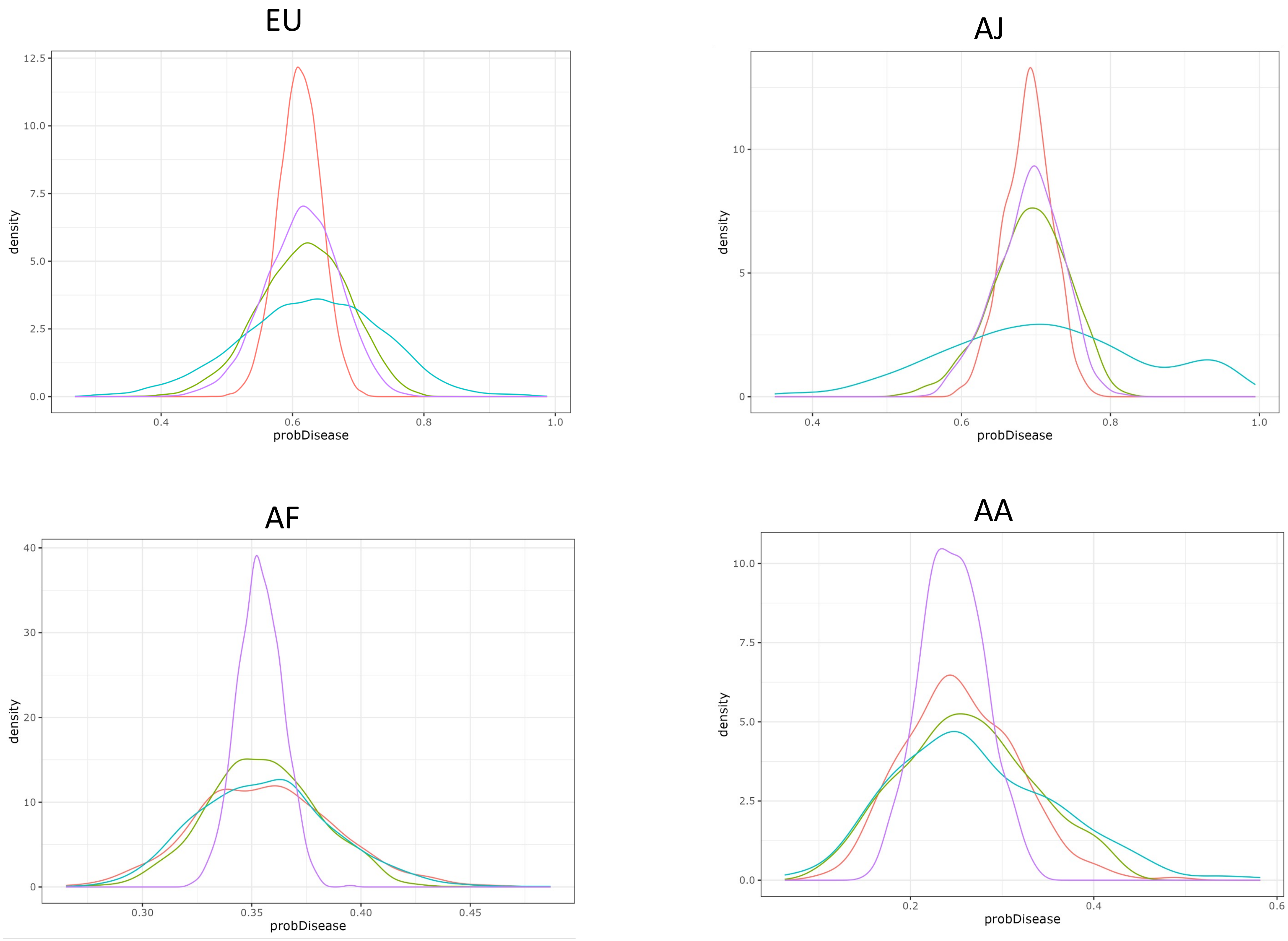


**Supplementary Figure 4a**: **Density plots comparison for each population.** Density plots comparing disease probabilities for each target data using four distinct summary statistics. Each panel represents a comparison of individual-level data. From left to right, the panels depict data from EUR, AFR, AJ, and AAC populations. The Y-axis represents density, while the X-axis denotes disease probability. Each distribution curve is color-coded: AAC is represented by pink, EAS by light green, AMR by purple, and EUR by calypso

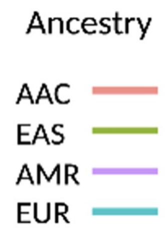

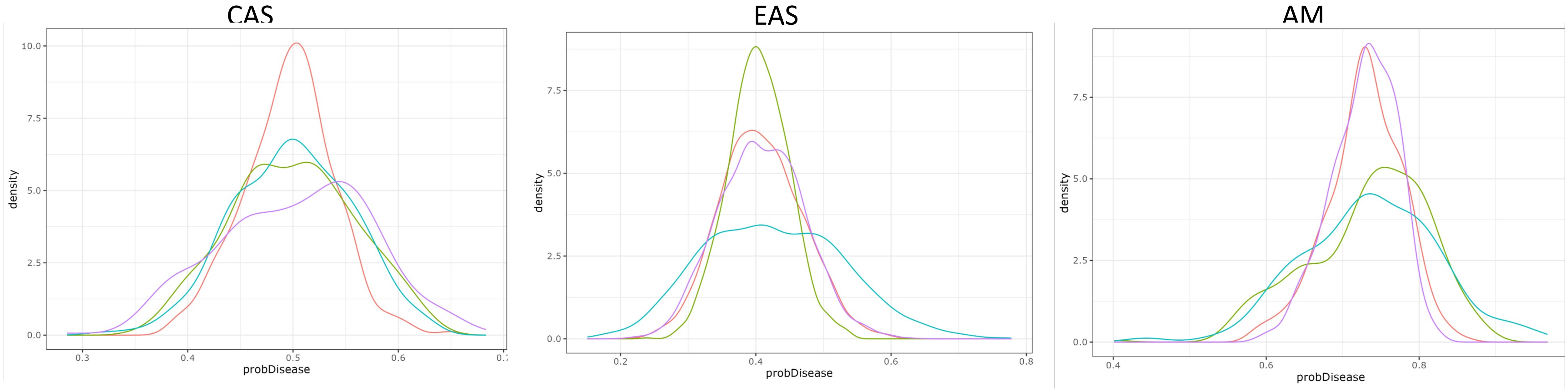


**Supplementary Figure 4b**: **Density plots comparison for each population.** Density plots comparing disease probabilities for each target data using four distinct summary statistics. Each panel represents a comparison of individual-level data. From left to right, the panels depict data from CAS, EAS, and EUR populations. The Y-axis represents density, while the X-axis denotes disease probability. Each distribution curve is color-coded: AAC is represented by pink, EAS by light green, AMR by purple, and EUR by calypso

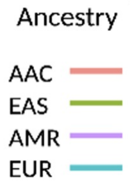

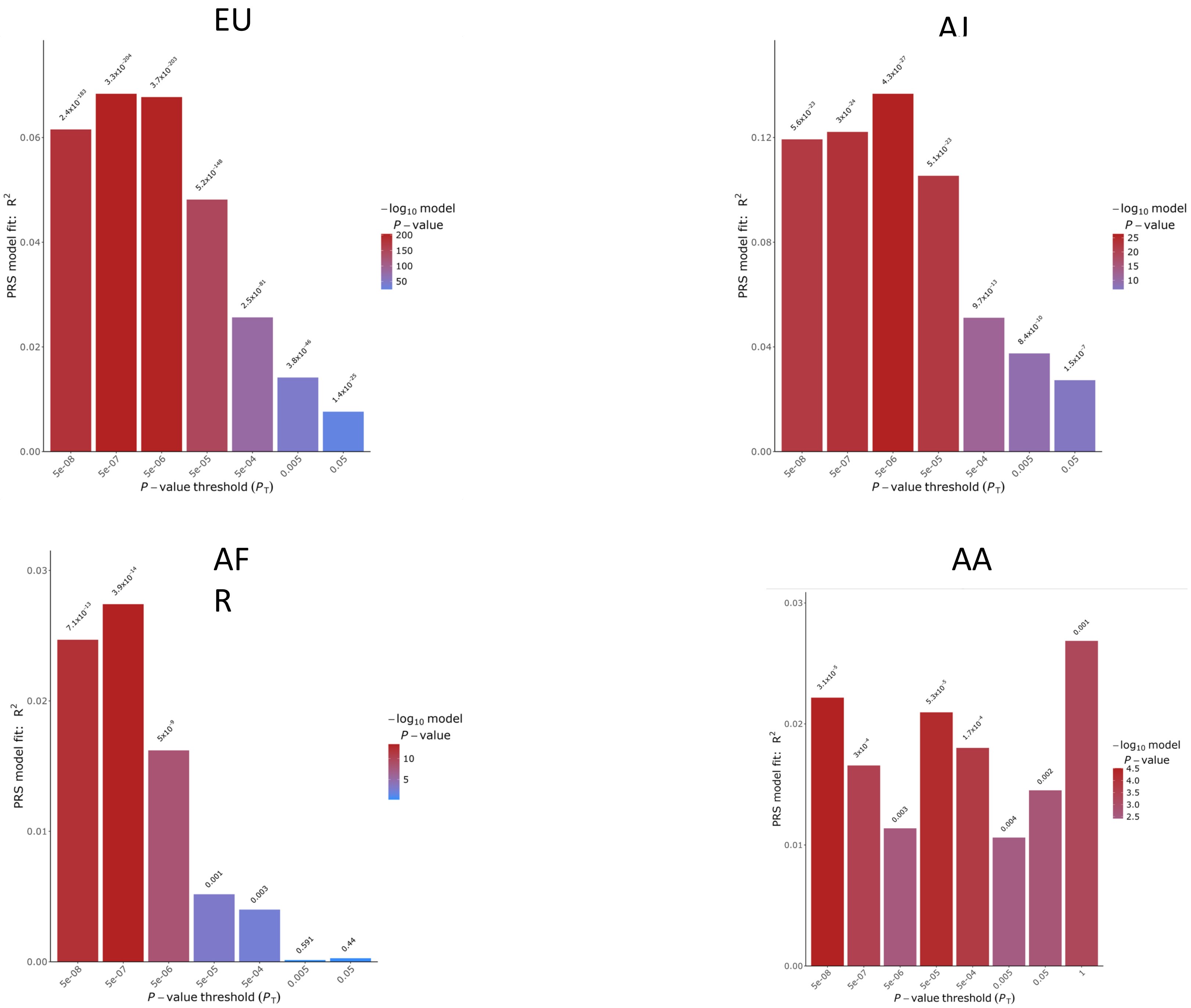


**Supplementary Figure 5a**: Multi-ancestry PRS bar plot representing an assessment of the PRS model's performance by p-value thresholding. The PRS Model Fit R^2 is plotted on the Y axis against the P-value Threshold on the X axis.

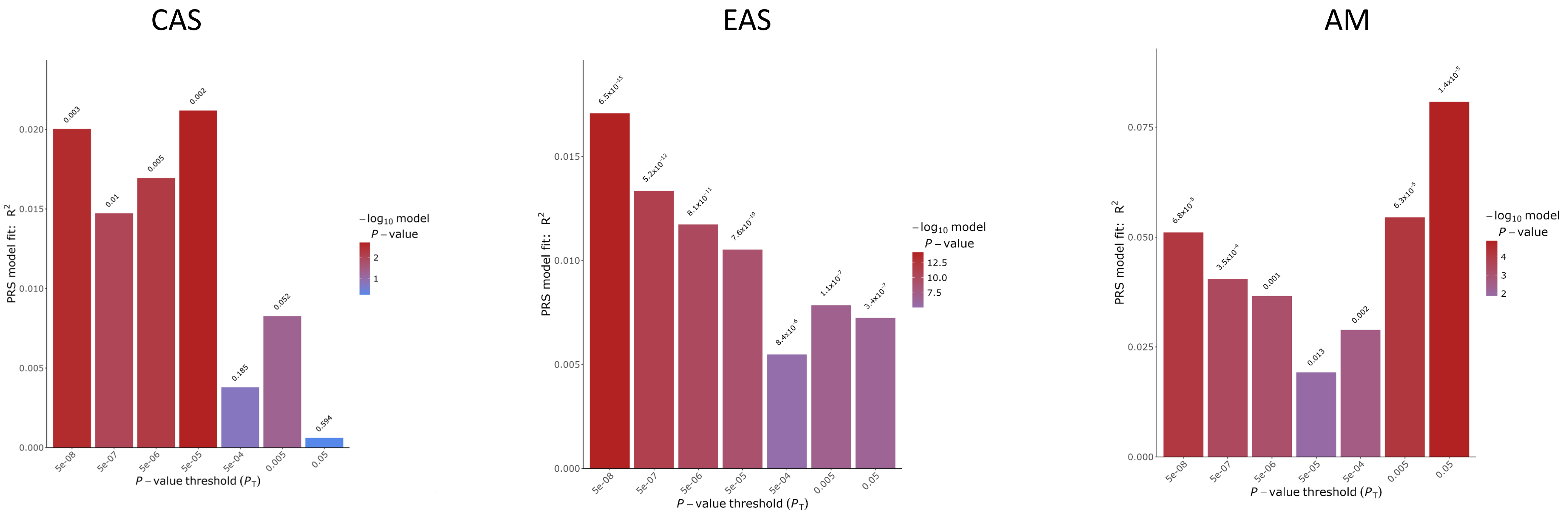


**Supplementary Figure 5b**: Multi-ancestry PRS bar plot representing an assessment of the PRS model's performance by p-value thresholding. The PRS Model Fit R^2 is plotted on the Y axis against the P-value Threshold on the X axis.
